# High ambient temperature during pregnancy and offspring cerebral palsy: A population-based study in California

**DOI:** 10.1101/2025.05.21.25328071

**Authors:** Haoran Zhuo, Joshua L. Warren, Giselle Bellia, Pin Wang, Kai Chen, Zeyan Liew, Tormod Rogne

## Abstract

**Background:** Current evidence on prenatal exposure to heat stress and childhood neurodevelopment is sparse. Cerebral palsy (CP) is the most common neuromotor disorder in childhood and there are growing concerns that environmental factors may play an etiological role. Our aim was to investigate whether prenatal exposure to high ambient temperature was associated with CP risk in the offspring.

**Methods:** We conducted a nested case-control study in California that included all CP cases identified from a statewide service system on developmental disabilities and randomly selected 20% of all live births without CP as controls during 2005-2015. Gestational weekly average temperatures were calculated from high resolution (1 km) daily mean temperature data based on maternal residential address. Extreme heat was defined as weekly averages above the 90^th^ percentile of the local temperature distribution. We implemented a distributed lag model within a logistic regression framework to estimate the associations between ambient temperatures increase, extreme heat and CP risk, across gestational week 0 to 31 covering early- and mid-pregnancy, and in the final seven weeks preceding birth capturing the late pregnancy. We also examined possible heterogeneity across maternal socio-demographic characteristics. Finally, we performed a sibling analysis to consider the influence of uncontrolled confounding.

**Findings:** The study population included 5,938 CP cases and 1,092,313 controls. There was an associated 2% increased odds of CP (95% credible interval: 1% to 5%) per 5 °C increase in ambient temperature in gestational week 0 to 3, and higher odds of 1.03 to 1.05 for extreme heat. A similar susceptible window in early pregnancy was observed in sibling analysis. We also observed a tendency of more pronounced associations for neighborhoods with higher social vulnerability and a cumulative association with higher temperature in the final seven weeks preceding births.

**Interpretation:** Early-pregnancy exposure to high ambient temperatures were associated with increased risk of childhood CP. While the estimated magnitude was small, our findings suggest that CP risk should be monitored in the population within the context of climate change.

**Funding:** Yale Planetary Solutions, National Institutes of Environmental Health Sciences

## Introduction

Rising ambient temperatures are increasingly recognized as a public health concern and are associated with adverse health outcomes in pregnant individuals and their developing fetuses.^1^ As a dynamic period, pregnancy is characterized by numerous tightly regulated physiological and psychological changes, rendering it particularly susceptible to heat stress.^2^ Evidence have suggested the prenatal origins of major childhood neurodevelopmental disorders, recognizing pregnancy period as a critical window for brain development.^3^ However, little is known regarding the impact of heat exposure during pregnancy on childhood neurodevelopment.

Cerebral palsy (CP) is a group of permanent nonprogressive motor and posture disorders that have clinical onsets in childhood.^4^ Most CP cases have prenatal origins with lesions occurring in the developing brain, while a clear etiology regarding what causes the brain damage remains largely unknown.^5^ Growing evidence has suggested environmental risks for CP, particularly prenatal exposures to environmental factors that could induce neuroinflammation or oxidative stress, which may further increase the susceptibility of fetal brain injury.^6–8^ Heat exposure during pregnancy, which can affect maternal physiological and cell responses that can lead to antenatal inflammation, infections, or placental insufficiency that increases the CP risk of offsprings.^9–11^ Additionally, heat stress has been shown to have short-term triggering effect on premature delivery, which is a key risk factor for CP.^12,13^

The aim of this population-based study in California was to evaluate whether prenatal exposure to high ambient temperature or extreme heat were associated with offspring CP. We also aimed to examine the relevant susceptible windows of exposures during pregnancy and possible heterogeneity across maternal socio-demographic characteristics. Finally, we assessed the influence from uncontrolled confounding bias through a sibling comparison design.

## Methods

The study protocol was approved by the official institutional review board at Yale University (IRB no. 2000028297) and the California Committee for the Protection of Human Subjects (project no. 12-10-0861) and was exempted from the informed consent requirement because there was no contact with human subjects. The study was reported by following the Strengthening and Reporting of Observational Studies in Epidemiology (STROBE) guidelines.

### Cases and controls ascertainment

We conducted a population-based case-control study of CP in California. CP cases were ascertained through a probabilistic linkage between the diagnostic records from California Department and Developmental Services (DDS) of 2005-2021 and the California birth records of 2005-2015. DDS is a state referral agency that provides diagnostic evaluations, services, and supports to all residents with developmental disabilities in California without financial or citizenship requirements through 21 statewide regional centers.^14^ CP cases in the DDS records was defined as a group of nonprogressive lesions or disorders in the brain characterized by abnormal movement or posture control that manifested in early childhood,^14^ having a high inter-rater reliability exceeding 0.85.^15^

The details of probabilistic linkage have been presented elsewhere.^7^ Briefly, we linked the California birth and the DDS records using personal identified information (e.g., name, date of birth, sex of child) through the LinkPlus software and crossed-checked less certain pairs by three research assistants, achieving the overall linkage rate of 93%. The population control group was a 20% randomly selected representative sample of all singleton births without CP in California of 2005-2015. We excluded all cases and controls with missing maternal residential address at delivery on the birth records, and gestational week data of <22 or >44 week that is susceptible to recording errors. The final sample for statistical analysis included 5,938 CP cases and 1,092,313 controls.

To assess the influence from uncontrolled confounding, we further utilized a sibling comparison design by identifying families with outcome-discordant siblings (i.e., families with both CP cases and non-CP sibling controls) from the entire California birth cohort, consisting of 1,758 CP cases and 2,316 non-CP sibling controls. The sibling identification was also based on a probability linkage approach similar as above, using linking variables of child’s last name, parental first name, last name, and date of birth.^16^

### Exposure assessment

The exposures of interest included high ambient temperature and extreme heat during pregnancy. Exposures to high ambient temperature was quantified by a continuous variable as per 5°C increase of the weekly-averages on daily mean temperatures, and extreme heat was defined binarily (e.g., Yes/No) as whether the weekly-averages of mean temperatures above the 90^th^ percentile (around 24.7 °C) of the distribution at each location during the study period of 2005-2015. To ascertain the exposures of interests, we geocoded the maternal residential addresses at delivery on point-address levels using the ArcGIS StreetMap Premium North America of 2013 release. Daily maximum and minimum air temperature at 2-meter readings with a spatial resolution of 1 km^2^ were obtained from the Daily Surface Weather Data for North America (Daymet, version 4; https://daymet.ornl.gov/). Daymet is a widely used model that follows strict testing procedure for uncertainty and provides accurate estimation of ambient temperature.^17^ Daily mean temperatures were calculated by averaging the daily maximum and minimum temperatures. Weekly estimates of ambient temperature and extreme heat were subsequently derived from the calculated daily means for gestational week 0 to 31 and for the final seven weeks preceding birth, based on the reported date of birth and last menstrual period documented in the birth records. We *a priori* focused on studying exposures in week 0 to 31 because approximately 30% of all CP cases are born preterm and the late gestational exposures are absent for these cases.^13^ To capture the exposure effects in late pregnancy, we examined exposures in the final seven weeks preceding birth.

### Statistical analyses

We performed a distributed lag model^18^ within a logistic regression framework to estimate the associations between ambient temperature and exposure to extreme heat by mutually considering exposures in gestational week 0 to 31 and the risk of CP. To capture the exposure in late pregnancy, we also perform a distributed lag model across weekly exposures in the final seven weeks preceding birth. We worked in the Bayesian setting for model fitting and modeled the distributed lag regression parameters using a Gaussian process prior distribution with time series correlation structure to address the potentially high correlation among weekly exposures, leading to more robust and smoothed estimates of susceptible windows of exposure.^18^ We assumed a linear relationship between weekly exposure and outcome for improved interpretability and visualization of susceptible windows (i.e., statistically significant gestational week-specific regression parameters), and also to more easily make inference on cumulative exposure effects (e.g., trimester). Potential non-linear exposure-outcome relationship was examined in sensitivity analyses. Full details regarding the statistical model, model fitting, and prior distributions w presented in Supplementary method. We also estimated the cumulative effects of exposures in the identified susceptible windows, within each trimester (week 0 to 13 as first trimester, week 14 to 28 as second trimester, and week 27 to 31 as early third trimester), and over the final seven weeks preceding birth.

All models were adjusted for a *priori* selected maternal, child, and community-level sociodemographic factors that might potentially confound the associations between temperature and CP, including: maternal age at delivery (≤18, 19–25, 26–30, 31–35, >35 years), self-reported race/ethnicity (African American or Black, Asian, Hispanic or Latinx of any race, non-Hispanic

White, others), self-reported education level (<12th grade, high school or diploma, college and above), primary payment source for prenatal care (government, private, other), parity (1, 2,≥3), and four domain-specific social vulnerability indexes (SVI, continuous)^19^ on census-tract level representing neighborhood socioeconomic status, household composition and disability, minority status and language, and housing type and transportation. We also adjusted for the child’s birth year (indicator for each year of 2005-2015) and season of conception (winter, spring, summer, fall) to address temporal confounding, and adjusted for birth county to address spatial confounding.

We conducted stratified subgroup analyses to investigate potential heterogeneity regarding CP subtypes (spastic, ataxic or dyskinetic) and socio-demographic characteristics including child’s sex (females, males), maternal race/ethnicity (Hispanic, non-Hispanic White; focused on these two due to statistical power), education level (high school and below, college and above), and census-tract level social vulnerability defined by the total SVI using the cutoffs of flag area from CDC definition (≥90^th^ percentile as high vulnerability, <90^th^ percentile as low vulnerability).^19^

Finally, we performed a sibling comparison study to assess the influence from uncontrolled confounding and investigated whether the associations between ambient temperature and extreme heat with CP risk holds. Sibling comparison analysis by design has controlled for all shared confounding factors among the siblings, regardless of whether they unmeasured or unknown.^20^ We implemented conditional logistic regression with strata on families for each gestational week separately from week 0 to 31, resulting in estimated OR and 95% confidence intervals for CP in each week without consideration of exposure in other weeks. Given that several familiar characteristics were comparable within siblings, we adjusted for time-varying factors including year of birth, maternal primary insurance type of prenatal care, age at delivery, and parity. We did not adjust for season of conception to maintain the exposure contrast at temporal level, given that most sibling pairs shared similar spatial locations.

### Sensitivity analyses

We conducted multiple sensitivity analyses to verify the robustness of our main findings. First, we extended the exposure windows to week 36 in our main model. Second, we tested the validity of the linear exposure-outcome relationship in the main models on ambient temperature by fitting a non-linear cubic spline of weekly exposures with degree freedoms of 3 to 6, focusing on the identified susceptible windows. Third, we assessed the heterogeneity in the estimated exposure effects during the final seven weeks preceding birth by the length of gestation into subgroups of 23-27, 28-31, 32-36, and 37-41. To address the potential collider bias in these stratified analyses,^21^ we implemented the fetus-at-risk approach where all ongoing pregnancy were included in the control group,^22^ for example, the control group of week 23-27 consists of non-CP births at week 23-27 and all ongoing pregnancy with gestational length of 28 weeks and longer.

## Results

Compared to non-CP controls, case mothers were more likely to give birth at an older age (>35 years old), to self-identified as Hispanic or Latinx and African American or Black, to have received lower educational attainment, and to utilize government insurance as their primary payment for prenatal care. Children with CP were more likely to be male and born at shorter gestational age (Table 1). The lowest and highest weekly averaged ambient temperature during pregnancy were −10.1°C and 39.9°C. The median and 90^th^ percentile of weekly temperature during the prenatal period were slightly higher among CP cases (17.5 °C and 24.9 °C) than controls (17.3 °C and 24.7 °C) (Supplementary Table 1).

**Table 1.** Characteristics of the case-control study sample in California, 2005–2015.

|  | Cerebral Palsy Cases<br>N=5 938 (%) | Controls<br>N=1 092 313 (%) |
| --- | --- | --- |
| Maternal age at delivery |  |  |
| ≤18 | 346 (5.8%) | 50,834 (4.7%) |
| 19-25 | 1,703 (28.7%) | 315,266 (28.9%) |
| 26-30 | 1,521 (25.6%) | 297,998 (27.3%) |
| 31-35 | 1,369 (23.1%) | 269,142 (24.6%) |
| >35 | 1,000 (16.8%) | 159,073 (14.6%) |
| Maternal race and ethnicity |  |  |
| Non-Hispanic White | 1,546 (26.0%) | 309,349 (28.3%) |
| Hispanic or Latinx of any race | 3,227 (54.3%) | 546,956 (50.1%) |
| African American or Black | 479 (8.1%) | 61,682 (5.6%) |
| Asian | 519 (8.7%) | 144,011 (13.2%) |
| Others (include Pacific Islanders and other) | 52 (0.9%) | 10,890 (1.0%) |
| Unknown | 115 (1.9%) | 19,425 (1.8%) |
| Maternal education level |  |  |
| <12th grade | 1,668 (28.1%) | 263,192 (24.1%) |
| High school or diploma | 2,844 (47.9%) | 495,362 (45.3%) |
| College and above | 1,175 (19.8%) | 288,749 (26.4%) |
| Unknown | 251 (4.2%) | 45,010 (4.1%) |
| Primary payment type for prenatal care |  |  |
| Government | 3,141 (52.9%) | 528,018 (48.3%) |
| Private | 2,537 (42.7%) | 516,241 (47.3%) |
| Other | 245 (4.2%) | 46,199 (4.2%) |
| Unknown | 15 (0.3%) | 1,855 (0.2%) |
| Sex of children |  |  |
| Male | 3,247 (54.7%) | 560,072 (51.3%) |
| Female | 2,691 (45.3%) | 532,241 (48.7%) |
| Parity |  |  |
| 1 | 2,388 (40.2%) | 426,803 (39.1%) |
| 2 | 1,696 (28.6%) | 344,810 (31.6%) |
| 3+ | 1,854 (31.2%) | 320,700 (29.4%) |
| Birth year |  |  |
| 2005-2010 | 3631 (61.2%) | 613,831 (56.2%) |
| 2011-2015 | 2307 (38.9%) | 478,482 (43.8%) |
| Season of conception |  |  |
| Spring (March-May) | 1,463 (24.6%) | 265,415 (24.3%) |
| Summer (June-August) | 1,425 (24.0%) | 266,129 (24.4%) |
| Fall (September-November) | 1,473 (24.8%) | 280,541 (25.7%) |
| Winter (December-February) | 1,577 (26.6%) | 280,228 (25.7%) |
| Gestational length (week) (mean ± SD) | 36.7 ± 4.9 | 39.1 ± 2.1 |
| Social vulnerability index (mean ± SD) |  |  |
| Total score | 0.62 ± 0.27 | 0.58 ± 0.28 |
| Socioeconomic domain | 0.62 ± 0.28 | 0.57 ± 0.29 |
| Household domain | 0.60 ± 0.27 | 0.56 ± 0.29 |
| Minority domain | 0.61 ± 0.27 | 0.58 ± 0.27 |
| Transportation domain | 0.57 ± 0.28 | 0.55 ± 0.28 |

We found that per 5 °C increase of temperature in each of the first four weeks during pregnancy there was an associated 2% increased odds of childhood CP (week 0, odds ratio, OR 1.02, 95% credible interval (CI) 0.99-1.05; week 1, OR 1.02, 95% CI 1.01 −1.05; week 2, OR 1.02, 95% CI 1.01-1.05; week 3, OR 1.02, 95% CI 1.00 −1.04) (Figure 1, Supplementary Table 2). Exposures to extreme heat in the same windows were associated with 3-5% increased odds of CP (Figure 1, Supplementary Table 2). The estimated cumulative associations over the first four weeks were 1.08 (95% CI 1.02 −1.15) for ambient temperature and 1.18 (95% CI 1.00 −1.43) for extreme heat (Table 2). There were positive associations between cumulative ambient temperature and extreme heat and the risk of CP in the first trimester, but these were less pronounced in the second trimester and were null in the early third trimester (Table 2) These results did not change when extended the window of assessment through gestational week 36 (Supplementary Figure 1).

**Figure 1.**
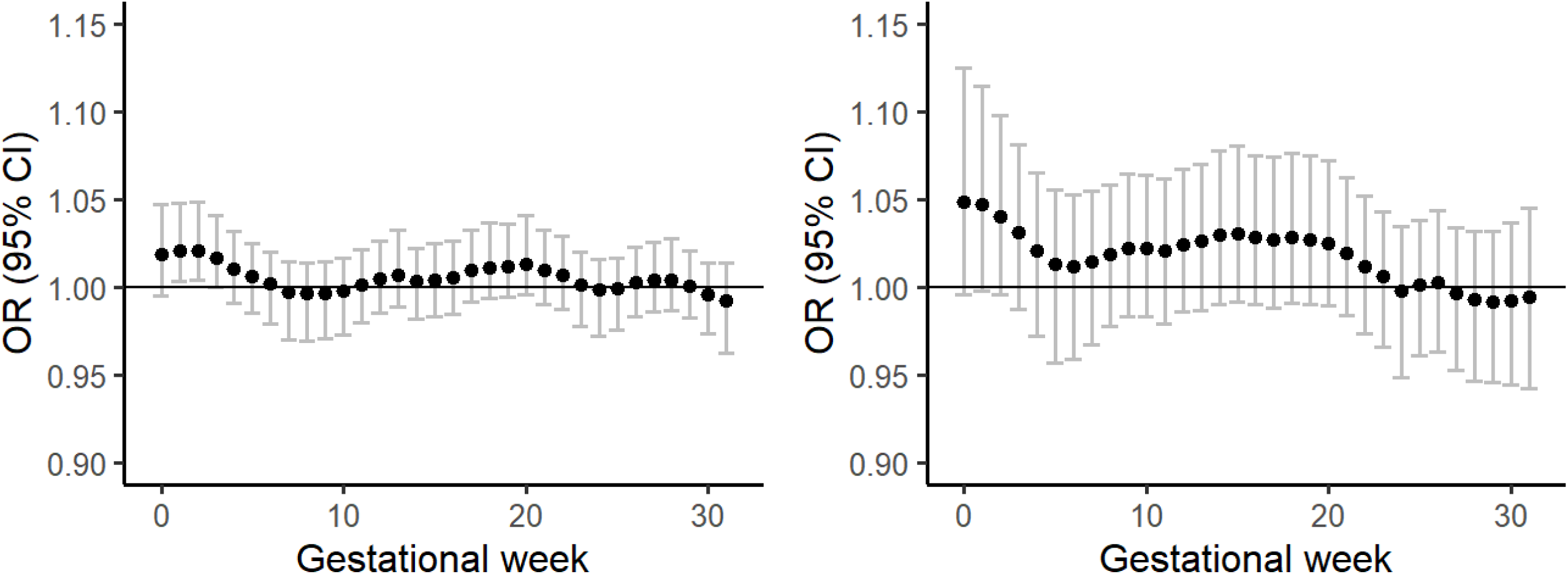
Associations between ambient temperature (left) and extreme heat (right) during pregnancy and the risk of cerebral palsy. Estimated odds ratio (ORs) and 95% credible intervals (CIs) of cerebral palsy for ambient temperature at per 5 °C increase and extreme heat defined as weekly mean temperature above the 90^th^ percentile in gestational weeks 0 to 31. Results were derived from the posterior distribution of regression parameters using a distributed lag logistic regression model fitted in the Bayesian setting, adjusted for year of birth, season of conception, birth county, maternal individual characteristics (race/ethnicity, education level, primary insurance type of prenatal care, age at delivery, parity), census-tract level social vulnerability (SES, household, transportation, minority).

**Table 2.** Cumulative associations between ambient temperature and extreme heat during pregnancy and the risk of cerebral palsy. Estimated cumulative odds ratio (ORs) and 95% credible intervals (CIs) of cerebral palsy for ambient temperature at per 5 °C increase and extreme heat defined as weekly mean temperature above the 90^th^ percentile over the identified susceptible windows (week 0 to 3), within each trimester, and in the final seven weeks preceding birth. Results were derived from the posterior distribution of regression parameters using a logistic-regression distributed lag logistic regression model fitted in the Bayesian setting, adjusted for year of birth, season of conception, birth county, maternal individual characteristics (race/ethnicity, education level, primary insurance type of prenatal care, age at delivery, parity), census-tract level social vulnerability (SES, household, transportation, minority).

| Exposure window | Ambient Temperature<br>OR (95% CI) | Extreme Heat<br>OR (95% CI) |
| --- | --- | --- |
| Susceptible window (week 0 to 3) | 1.08 (1.02, 1.15) | 1.18 (1.00, 1.43) |
| Trimester 1 (week 0 to 13) | 1.10 (1.03, 1.17) | 1.43 (1.00, 2.05) |
| Trimester 2 (week 14 to 27) | 1.08 (1.01, 1.16) | 1.25 (0.93, 1.70) |
| Trimester 3 (week 28 to 31) | 0.99 (0.94, 1.04) | 0.97 (0.83, 1.13) |
| Final seven weeks preceding birth | 1.05 (1.01, 1.09) | 0.95 (0.78, 1.16) |

Neither exposures to higher ambient temperature nor extreme heat in each week of the final seven weeks preceding birth were associated with higher risk of CP (Supplementary Figure 2), while the cumulative effects of higher ambient temperature over the final seven weeks preceding birth had an associated 5% increased odds of CP (Table 2) and the estimated effects were pronounced among those born extremely preterm (Supplementary Figure 3).

Similar susceptible window of exposures during pregnancy in week 0 to 3 were identified across stratified subgroup analyses although with lower precision (Supplementary Figure 4 to Supplementary Figure 8). For the cumulative effects of exposures over this period, we observed slightly stronger point estimates among males, spastic CP subtypes, and mothers who lived in neighborhoods with high social vulnerability, while their confidence intervals were largely overlapped across the subgroup strata (Table 3). Stronger estimated effects under exposures to extreme heat, but not high ambient temperature, were also observed among Non-Hispanic White mothers and those with high education level of college and above (Table 3).

**Table 3.** Stratified analyses on cumulative associations between ambient temperature and extreme heat during pregnancy and the risk of cerebral palsy. All p-values for interactions were larger than 0.05. Estimated cumulative odds ratio (ORs) and 95% credible intervals (CIs) of cerebral palsy for ambient temperature at per 5 °C increase and extreme heat defined as weekly mean temperature above the 90^th^ percentile over the identified susceptible windows of gestational week 0 to 3. Results were derived from the posterior distribution of regression parameters using a logistic-regression distributed lag logistic regression model fitted in the Bayesian setting, adjusted for year of birth, season of conception, birth county, and covariates in the following except for the stratified items: maternal individual characteristics (race/ethnicity, education level, primary insurance type of prenatal care, age at delivery, parity), and census-tract level total social vulnerability).

| Cumulative associations in gestational week 0 to 3 | Ambient Temperature<br>OR (95% CI) | Extreme Heat<br>OR (95% CI) |
| --- | --- | --- |
| **Main analysis** |  |  |
| Overall case-control sample | 1.08 (1.02, 1.15) | 1.18 (1.00, 1.43) |
| **Stratified analyses** |  |  |
| Cerebral palsy subtypes |  |  |
| Spastic | 1.13 (1.04, 1.24) | 1.07 (0.86, 1.33) |
| Ataxic & Dyskinetic | 1.02 (0.88, 1.17) | 1.67 (0.94, 3.18) |
| Sex of child |  |  |
| Male | 1.12 (1.04, 1.23) | 1.30 (1.01, 1.71) |
| Female | 1.05 (0.99, 1.13) | 1.11 (0.88, 1.43) |
| Maternal race/ethnicity |  |  |
| Hispanic | 1.06 (1.01, 1.15) | 1.15 (0.92, 1.45) |
| Non-Hispanic White | 1.09 (1.00, 1.22) | 1.55 (1.07, 2.28) |
| Maternal education level |  |  |
| High school and lower | 1.07 (1.01, 1.15) | 1.15 (0.94, 1.44) |
| College and above | 1.06 (0.98, 1.16) | 1.46 (1.02, 2.17) |
| Census-tract level social vulnerability |  |  |
| High (SVI≥90<sup>th</sup> percentile) | 1.07 (1.01, 1.15) | 1.30 (1.01, 1.72) |
| Low (SVI <90<sup>th</sup> percentile) | 1.10 (1.01, 1.22) | 1.11 (0.85, 1.43) |

Our sensitivity analyses on the potential non-linear relationship of ambient temperature in week 0 to 3 on the risk CP did not indicate evident non-linearity (Supplementary Figure 9). As expected in the sibling comparison design, several time-invariant maternal characteristics such as maternal race/ethnicity, educational level, and neighborhood SVI were highly comparable among CP cases and their sibling controls among the outcome-discordant siblings (Supplementary Table 3). By using siblings with more comparable familiar characteristics, we identified similar susceptible windows of exposures to high ambient temperature and extreme heat in early pregnancy covering week 0 to 3 (Figure 2, Supplementary Table 4).

**Figure 2.**
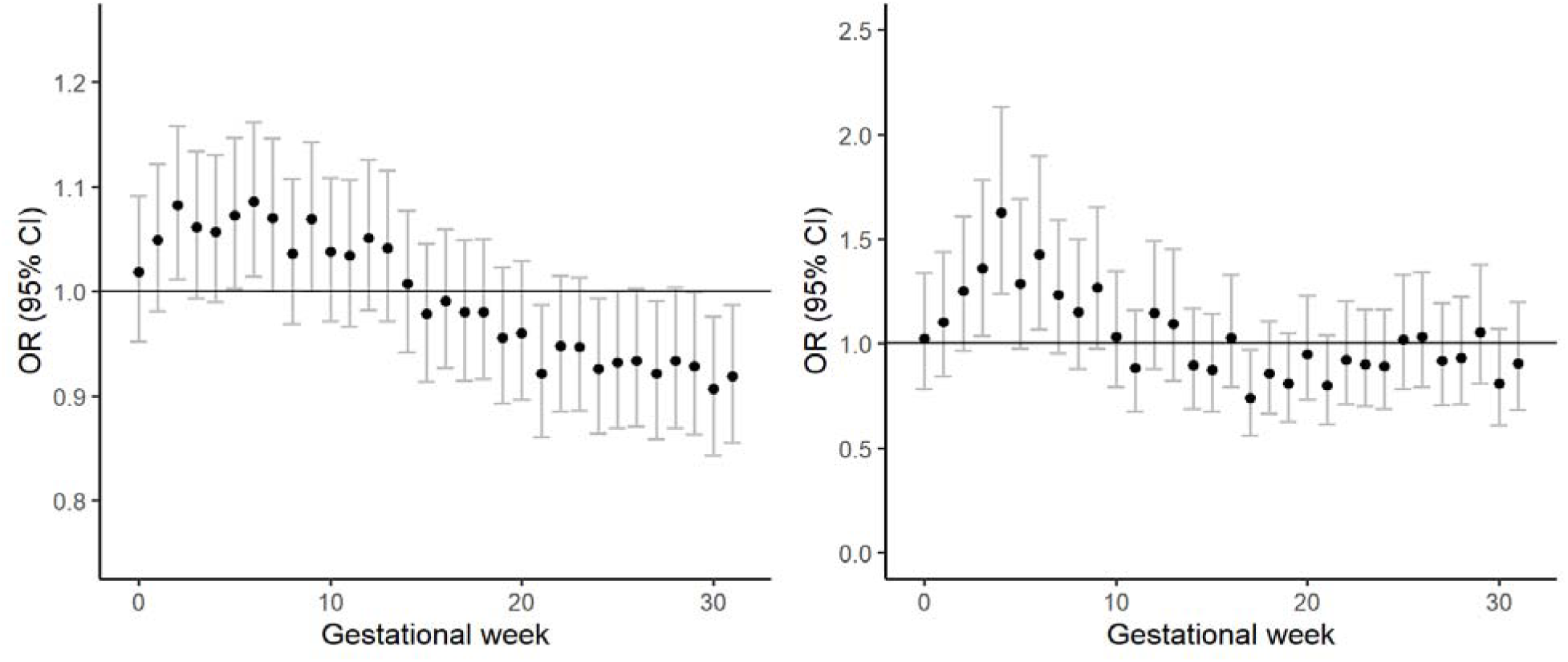
Associations between ambient temperature (left) and extreme heat (right) during pregnancy and the risk of cerebral palsy among outcome-discordant siblings. Estimated odds ratio (ORs) and 95% credible intervals (CIs) of cerebral palsy for ambient temperature at per 5 °C increase and extreme heat defined as weekly mean temperature above the 90^th^ percentile in gestational weeks 0 to 31. Results were derived from conditional logistic regression applied separately to each gestational week, adjusted for year of birth, maternal primary insurance type of prenatal care, age at delivery, and parity.

## Discussion

In this population-based study of CP in California, we observed an elevated risk of childhood CP under prenatal exposures to higher ambient temperature, with the most susceptible window in early pregnancy covering gestational weeks 0 to 3. These findings remained robust across a series of analyses considering different temperature exposure matrices, varying exposure window lengths, and comprehensive confounding assessments. Additionally, a positive trend of higher CP risk was observed under cumulative exposure to elevated ambient temperature over the first trimester, the second trimester, and the final seven weeks preceding birth.

The biological mechanisms linking high ambient temperature and CP risk are largely unknown and several hypothesized pathways may underlie the observed associations (Supplementary Figure 10). Prenatal exposure to high temperature involves both maternal physiological responses and cell responses that can either directly affect fetal brain development or indirectly induce prenatal or perinatal risk factors of CP. ^9–11^ Specifically, prenatal exposure to high temperature can induce excessive maternal sweating and dehydration, leading to reduced uterine blood flow and diminished oxygen delivery to the fetal brain.^9^ In addition, high temperatures can disable the protective effects of heat shock proteins on other essential proteins, such as hemoglobin that carries oxygen to the fetal brain.^10^ Together, these mechanisms can directly affect fetal brain malformations or alter brain structure in early pregnancy. Secondly, heat stress can induce breakdowns of heat shock proteins, leading to systematic inflammation and oxidative stress that triggers preterm birth,^11^ a well-known risk factor for CP,^13^ contributing to higher susceptibility of CP development in exposure window of preceding birth. Thirdly, normal physiological responses to high ambient temperatures may become exaggerated during pregnancy, increasing the susceptibility of CP by inducing pregnancy complications, restricted fetal growth, and antenatal infections that happen in mid to late pregnancy.^23,24^ Given that CP is a complex spectrum of various subtypes with multifactorial etiology, prenatal exposure to high ambient temperature at different stage of pregnancy may influence CP risk through multiple biological pathways, although findings from our study suggested the most susceptible windows were in the early pregnancy.

Although current evidence on perinatal exposure to ambient temperature and offspring neurodevelopment remains limited, exposures occurring in early gestation period (including pre-conception) have been more frequently suggested as susceptible windows, ^25,26^ showing consistency with our findings on CP. In previous studies, childhood neurodevelopmental delays (including motor function) have been associated with prenatal exposure to heatwaves in first trimester of pregnancy in a Chinese cohort study.^25^ Another US case-control study found that early pregnancy exposure to heatwaves, covering a period from eight weeks pre-conception to four weeks post-conception, was associated with offspring neural tube defects.^26^ In fact, early pregnancy establishes the fundamental brain structures and has been suggested to have heightened vulnerability of CP risk under exposures to various other environmental disruptors, such as toxic air contaminants^8^ and pesticides.^6^ Emerging evidence also recognized their interactions with high ambient temperature, suggesting an amplified neurotoxicity and developmental toxicity from the co-existence of hot weather and ambient pollutants^27^ that may further increase CP risk. In addition, studies have observed a consistent short-term triggering effect of extreme heat in late pregnancy (or weeks preceding birth) on preterm birth,^12^ a key risk factor for CP, but it only explains less than 20% of all cases.^13^ Evidence also observed a small amount (<8%) of overall effect from seasonally varied environmental factors to childhood CP was mediated through preterm birth.^7^ In together, these indicated that late-pregnancy triggering effect of high ambient temperature may play limited role in CP development. Finally, we observed a cumulative association with high ambient temperature in the second trimester, which might be explained by several perinatal risk factors of CP that happen in mid- or early-late pregnancy such as maternal pregnancy complications and neonatal adverse birth outcomes.^1^ However, no clear evidence regarding the susceptible window of heat exposure on these perinatal factors and requires further investigation.

Our stratified analyses did not support strong sociodemographic disparity over the magnitude of estimated CP risk under prenatal exposure to high ambient temperature. A slightly stronger association was observed among more socially vulnerable neighborhoods, suggesting a potential interplay between ambient temperature and other neighborhood-level social and environmental factors. A smaller magnitude of increased risk of CP were observed among individuals who were racially or socially more vulnerable. One possible explanation of this counter intuitive observation could be that the low SES group have higher rate of miscarriage and infant mortality, which in turn resulting in larger live birth bias that hinder the identification of harmful effects of environmental factors.^28^ Future research regarding the complex interplay among environmental, social, and socio-environmental factors at both individual and neighborhood levels is warranted to understand the multifactorial etiology of CP in the context of changing climate.

### Strengths and limitations

Our study is one of the largest population-based studies on CP. The registry-based linkage design eliminated the risk of recall bias and self-selection bias. In addition, we conducted a robust control for confounding factors, including shared unmeasured confounding in the family through sibling comparison design.

Nevertheless, there are also several limitations that should be noted. First, ambient temperature at maternal residential address may not reflect individual-level exposure to heat, which can be modified by individual behaviors, access to air conditioning, and living conditions. Furthermore, while daily mean temperature reflects the overall temperature experienced throughout the day, it may not capture other aspects of temperature exposure such as nighttime exposure or temperature variability. Exposure misclassification cannot be ruled out from the residential mobility during pregnancy. While 9%–32% of families are estimated to have moved during pregnancy, and most moving happened locally within 10 km,^29^ which is unlikely to have distinct climate conditions. Second, there is a possibility of outcome misclassification given that DDS may not capture mild CP cases that are in less need of services. However, this number is likely to be small given the rarity of CP prevalence. Additionally, this misclassification would likely be nondifferential to ambient temperature and thereby bias the estimate towards the null. Third, our study was restricted to live births, which makes our results susceptible to live-birth bias. That said, simulation studies have shown that when an environmental exposure differentially affects fetal losses among pregnancies of those susceptible to childhood health outcomes – which may be the case in the context of temperature and CP – the bias tends to be towards the null and may hinder the identification of exposure-outcome disparities.^28,30^ Finally, the current version of Bayesian distributed lag model was not able to handle matched data structure, so the sibling analyses were run by fitting separate conditional logistic regressions without mutually adjusted for each week and the estimated effects might not be directly comparable to main models.

Additionally, season of conception was not adjusted in the sibling analyses to maintain exposure contrast at temporal level, which may leave the potentiality of seasonal trend of exposure in the model.

## Conclusion

Our study suggested that exposure to high ambient temperatures in early pregnancy may play a role in CP etiology, which offers important insights regarding the future disease burden of childhood neurodevelopmental disorders in the context of climate change.

## Data Availability

The data that support findings from this study are from the California Department of Public Health and Department of Developmental Services. The authors have no rights to share the data.

