## Supplementary Materials for "High ambient temperature during pregnancy and offspring cerebral palsy: A population-based study in California"

**Supplementary Methods**

To identify susceptible window of exposure to ambient temperature during pregnancy, we implemented a Bayesian distributed lag model with logistic regression by utilizing the "GPCW" R package. Our fitted model was modified on the originally developed spatial-temporal hierarchical Bayesian logistic regression to estimate critical windows of susceptibility,[1] by leaving out the spatial aspect of the original model. This method is relied on a temporally structured Gaussian process to mutually consider exposures in all susceptible windows of gestational week and their lag effects. The model fitting utilized Markov chain Monte Carlo (MCMC) sampling techniques. From the sampled posterior distributions, we calculated the point estimates of exposure effect from the posterior means and derived the 95% quantile-based credible intervals (compared to confidence intervals in the frequentist analysis setting) to quantify the uncertainty of the parameters.

The fitted Bayesian Gaussian Process Model for Critical Window (GPCW) was as the following:

$$Y_i \mid \boldsymbol{\beta},\boldsymbol{\theta} \overset{\text{ind}}{\sim} \text{Binomial}\,\{p_i(\boldsymbol{\beta},\boldsymbol{\theta})\},\ i = 1, \ldots, \text{n};$$

$$\log\left\{\frac{p_i(\boldsymbol{\beta},\boldsymbol{\theta})}{1 - p_i(\boldsymbol{\beta},\boldsymbol{\theta})}\right\} = \mathbf{x}_i^{\text{T}}\boldsymbol{\beta} + \sum_{j=1}^{m_i} \text{z}_{ij}\,\theta(j);$$

$$\boldsymbol{\theta} = \{\theta\,(1), \ldots, \theta\,(\text{m})\}^{\text{T}} \mid \sigma_\theta^2, \phi \sim \text{MVN}\{\mathbf{0}_m, \sigma_\theta^2\,\Sigma(\phi)\}$$

$$\Sigma(\phi)_{ij} = \exp\{-\phi|i - j|\},\ \phi > 0$$

Input variables

- $Y_i$ : Outcome variable for individual *i*, the total number of study population is n
- $z_{ij}$ : Exposure for individual *i* at week *j;* the total number of gestational week is m
- $\mathbf{x}_i$ : Covariates matrix for individual *i*

Parameters

- $\theta(\text{j})$ : Coefficient for exposure at week *j,* modeled using a Gaussian process prior distribution (MVN) with mean zero and covariance matrix $\sigma_\theta^2\,\Sigma(\phi)$.
- $\boldsymbol{\beta}$: Coefficient matrix for all covariates, include intercepts and adjusted covariates in the model. We assumed the prior distribution were: $\beta_{1,..,k} \overset{\text{iid}}{\sim} N(0, \sigma_\beta^2)$ with *k* represents the number of covariates and $\sigma_\beta^2$ = 10,000.
- $\sigma_\theta^2\,\Sigma(\phi)$ : Covariance matrix for exposure assumed that pollutants in closer time window are more correlated, this allows the solution to potential multicollinearity issue in the traditional way of co-adjusting for multiple exposures in the same model. We assumed the prior distributions were: $\sigma_\theta^2 \sim$ Inverse Gamma ($a_{\sigma_\theta^2}$, $b_{\sigma_\theta^2}$) with default values of $a_{\sigma_\theta^2}$ =3 and

$b_{\sigma_\theta^2}$ =2, $\phi \sim$ Uniform ($a_\phi$ , $b_\phi$ ) with default values of $a_\phi$ = log(0.9999) / {- (m-1)}, $b_\phi$ = log(0.0001)/(-1).

Model fitting

For exposures to ambient temperature at per 5°C increase, we created a standardized exposure matrix by subtracting the median values and divided by five. For individuals with shorter gestational length than 32 weeks (or have any missing values before and at week 31), we replaced their missing values with the median values in the exposure matrix before standardization in order to run the Bayesian distributed lag model. For covariates matrix, we standardized continuous variables (e.g. social vulnerability index) and created indicator variables for each category of the categorical variable (e.g., separate indicators for Hispanic, Non-Hispanic White, African American/Non-Hispanic Black, Asian, and Other race/ethnicity). We implemented 100,000 times MCMC sample with 50,000 times burn-in period using above specified model and prior distribution.

### Supplementary Results

**Supplementary Table 1.** Temperature distribution among cerebral palsy cases and controls

**Supplementary Table 2.** Estimated odds ratio (ORs) and 95% credible intervals (CIs) for cerebral palsy exposed to ambient temperature and extreme heat in gestational week 0 to 31 during pregnancy (match Figure 1).

**Supplementary Table 3.** Characteristics of outcome-discordant siblings in California birth cohort, 2007–2015.

**Supplementary Table 4.** Estimated odds ratio (ORs) and 95% confidence intervals (CIs) for the cerebral palsy exposed to ambient temperature and extreme heat in gestational week 0 to 31 during pregnancy among siblings (match Figure 2).

**Supplementary Figure 1**. Estimated odds ratio (ORs) and 95% credible intervals (CIs) for the cerebral palsy exposed to ambient temperature and extreme heat in gestational week 0 to 36 during pregnancy.

**Supplementary Figure 2.** Estimated odds ratio (ORs) and 95% credible intervals (CIs) for the cerebral palsy exposed to ambient temperature and extreme heat in the last seven weeks preceding birth.

**Supplementary Figure 3.** Cumulative Odds Ratios (ORs) and 95% CIs for cerebral palsy according to exposures to ambient temperature and extreme heat in last seven weeks preceding birth, stratified by length of gestation.

**Supplementary Figure 4.** Estimated odds ratio (ORs) and 95% credible intervals (CIs) for the cerebral palsy exposed to ambient temperature and extreme heat in gestational week 0 to 31 during pregnancy, stratified by child's sex.

**Supplementary Figure 5.** Estimated odds ratio (ORs) and 95% credible intervals (CIs) for the cerebral palsy exposed to ambient temperature and extreme heat in gestational week 0 to 31 during pregnancy, stratified by cerebral palsy subtypes.

**Supplementary Figure 6.** Estimated odds ratio (ORs) and 95% credible intervals (CIs) for the cerebral palsy exposed to ambient temperature and extreme heat in gestational week 0 to 31 during pregnancy, stratified by maternal race/ethnicity.

**Supplementary Figure 7.** Estimated odds ratio (ORs) and 95% credible intervals (CIs) for the cerebral palsy exposed to ambient temperature and extreme heat in gestational week 0 to 31 during pregnancy, stratified by maternal education level.

**Supplementary Figure 8.** Estimated odds ratio (ORs) and 95% credible intervals (CIs) for the cerebral palsy exposed to ambient temperature and extreme heat in gestational week 0 to 31 during pregnancy, stratified by census-tract level social vulnerability index.

**Supplementary Figure 9.** Non-linear relationship of ambient temperature and cerebral palsy in gestational week 0 to 3, with degree freedom ranging 3 to 6.

**Supplementary Figure 10.** Underlying biological pathways from ambient temperature exposure and cerebral palsy.

**Supplementary Table 1. Temperature distribution among cerebral palsy cases and controls**

| Gestational week | Controls | | | | Cerebral Palsy Cases | | | |
|---|---|---|---|---|---|---|---|---|
| | N | Median ± SD | Min | Max | N | Median ± SD | Min | Max |
| week0 | 1092313 | 17.27 ± 5.31 | -9.14 | 39.37 | 5938 | 17.35 ± 5.36 | -3.14 | 37.18 |
| week1 | 1092313 | 17.22 ± 5.32 | -9.63 | 39.18 | 5938 | 17.25 ± 5.41 | -5.44 | 37.33 |
| week2 | 1092313 | 17.22 ± 5.33 | -10.02 | 39.2 | 5938 | 17.36 ± 5.41 | -4.62 | 35.22 |
| week3 | 1092313 | 17.20 ± 5.31 | -9.88 | 39.16 | 5938 | 17.38 ± 5.34 | 0.4 | 36.47 |
| week4 | 1092313 | 17.23 ± 5.33 | -9.21 | 39.18 | 5938 | 17.40 ± 5.36 | -0.52 | 37.32 |
| week5 | 1092313 | 17.18 ± 5.33 | -9.95 | 39.17 | 5938 | 17.25 ± 5.38 | -1.81 | 36.49 |
| week6 | 1092313 | 17.16 ± 5.34 | -10.02 | 39.2 | 5938 | 17.31 ± 5.38 | -1.87 | 37.3 |
| week7 | 1092313 | 17.16 ± 5.33 | -9.33 | 38.94 | 5938 | 17.37 ± 5.36 | -2.49 | 36.43 |
| week8 | 1092313 | 17.19 ± 5.33 | -8.5 | 39.58 | 5938 | 17.29 ± 5.36 | -3.02 | 37.69 |
| week9 | 1092313 | 17.16 ± 5.34 | -9.4 | 39.37 | 5938 | 17.30 ± 5.39 | -1.02 | 37.26 |
| week10 | 1092313 | 17.15 ± 5.35 | -8.05 | 39.09 | 5938 | 17.31 ± 5.40 | -3.67 | 36.07 |
| week11 | 1092313 | 17.18 ± 5.34 | -8.02 | 39.25 | 5938 | 17.31 ± 5.38 | -3.84 | 35.15 |
| week12 | 1092313 | 17.20 ± 5.31 | -9.02 | 39.91 | 5938 | 17.40 ± 5.35 | -1.57 | 37.88 |
| week13 | 1092313 | 17.25 ± 5.33 | -8.61 | 38.94 | 5938 | 17.47 ± 5.31 | -1.99 | 37.14 |
| week14 | 1092313 | 17.21 ± 5.33 | -7.86 | 39.61 | 5938 | 17.39 ± 5.37 | -7.45 | 36.26 |
| week15 | 1092313 | 17.25 ± 5.34 | -8.27 | 39.25 | 5938 | 17.53 ± 5.37 | -2.96 | 35.35 |
| week16 | 1092313 | 17.25 ± 5.31 | -9.07 | 39.18 | 5938 | 17.45 ± 5.33 | -6.33 | 36.76 |
| week17 | 1092313 | 17.29 ± 5.32 | -9.34 | 39.27 | 5938 | 17.68 ± 5.32 | -1.2 | 37.73 |
| week18 | 1092313 | 17.29 ± 5.33 | -8.23 | 39.14 | 5938 | 17.56 ± 5.30 | 0.44 | 37.2 |
| week19 | 1092313 | 17.31 ± 5.34 | -9.74 | 38.85 | 5938 | 17.48 ± 5.32 | -0.06 | 36.04 |
| week20 | 1092313 | 17.33 ± 5.33 | -7.73 | 38.85 | 5938 | 17.67 ± 5.34 | -4.25 | 36.81 |
| week21 | 1092313 | 17.39 ± 5.32 | -8 | 39.17 | 5938 | 17.59 ± 5.30 | -0.96 | 37.1 |
| week22 | 1092313 | 17.39 ± 5.34 | -8.67 | 39.18 | 5938 | 17.66 ± 5.35 | -0.62 | 38.77 |
| week23 | 1091941 | 17.39 ± 5.35 | -8.5 | 39.46 | 5890 | 17.63 ± 5.38 | -1.66 | 37.65 |
| week24 | 1091225 | 17.43 ± 5.35 | -9.44 | 38.93 | 5756 | 17.61 ± 5.36 | -5.35 | 37.13 |
| week25 | 1090370 | 17.44 ± 5.34 | -8.56 | 39.08 | 5604 | 17.66 ± 5.36 | -4.6 | 35.66 |
| week26 | 1089357 | 17.51 ± 5.37 | -9.88 | 39.18 | 5472 | 17.71 ± 5.42 | -4.39 | 36.64 |
| week27 | 1088098 | 17.48 ± 5.40 | -10.12 | 39.69 | 5340 | 17.74 ± 5.41 | -3.07 | 37.04 |
| week28 | 1086503 | 17.52 ± 5.42 | -9.86 | 39.35 | 5217 | 17.82 ± 5.43 | -1.4 | 36.85 |
| week29 | 1084564 | 17.51 ± 5.39 | -8.89 | 39.1 | 5101 | 17.69 ± 5.40 | -7.67 | 38.05 |
| week30 | 1081994 | 17.58 ± 5.41 | -9.11 | 39.18 | 4994 | 17.69 ± 5.46 | -6.64 | 37.02 |
| week31 | 1078511 | 17.59 ± 5.42 | -9.21 | 39.43 | 4896 | 17.67 ± 5.48 | -4.62 | 37.1 |
| week32 | 1073355 | 17.59 ± 5.43 | -8.25 | 38.81 | 4792 | 17.66 ± 5.43 | -4.43 | 37.64 |
| week33 | 1065707 | 17.60 ± 5.41 | -8.82 | 39.05 | 4665 | 17.79 ± 5.42 | -5.82 | 36.89 |
| week34 | 1052150 | 17.62 ± 5.41 | -9.12 | 39.14 | 4505 | 17.69 ± 5.38 | -6.04 | 38.01 |
| week35 | 1030699 | 17.63 ± 5.42 | -9.05 | 38.97 | 4322 | 17.91 ± 5.45 | -1.02 | 36.37 |
| week36 | 991978 | 17.60 ± 5.42 | -9.62 | 39.16 | 4006 | 17.79 ± 5.54 | -2.4 | 37.23 |
| week37 | 909051 | 17.60 ± 5.41 | -7.62 | 39 | 3516 | 17.68 ± 5.48 | -3.71 | 36.9 |
| week38 | 732172 | 17.56 ± 5.39 | -8.79 | 39.17 | 2723 | 17.67 ± 5.35 | 1.17 | 36.95 |
| week39 | 422333 | 17.50 ± 5.41 | -6.73 | 38.72 | 1607 | 17.48 ± 5.54 | -1.01 | 35.62 |
| week40 | 173204 | 17.33 ± 5.49 | -6.76 | 38.35 | 704 | 17.02 ± 5.81 | -3.48 | 35.1 |
| week41 | 53511 | 17.20 ± 5.62 | -7.54 | 38.62 | 254 | 17.25 ± 5.72 | 2.48 | 31.7 |

**Supplementary Table 2. Estimated odds ratio (ORs) and 95% credible intervals (CIs) for cerebral palsy exposed to ambient temperature and extreme heat (match Figure 1)**

| Gestational Week | Ambient Temperature | | Extreme heat (>90th percentile) | |
|---|---|---|---|---|
| | OR | 95% CI | OR | 95% CI |
| week0 | 1.02 | (0.99, 1.05) | 1.05 | (1.00, 1.13) |
| week1 | 1.02 | (1.01, 1.05) | 1.05 | (1.00, 1.12) |
| week2 | 1.02 | (1.01, 1.05) | 1.04 | (1.00, 1.10) |
| week3 | 1.02 | (1.00, 1.04) | 1.03 | (0.99, 1.08) |
| week4 | 1.01 | (0.99, 1.03) | 1.02 | (0.97, 1.07) |
| week5 | 1.01 | (0.99, 1.02) | 1.01 | (0.96, 1.05) |
| week6 | 1.00 | (0.98, 1.02) | 1.01 | (0.96, 1.05) |
| week7 | 1.00 | (0.97, 1.01) | 1.01 | (0.97, 1.05) |
| week8 | 1.00 | (0.97, 1.01) | 1.02 | (0.97, 1.06) |
| week9 | 1.00 | (0.97, 1.01) | 1.02 | (0.98, 1.06) |
| week10 | 1.00 | (0.97, 1.02) | 1.02 | (0.98, 1.07) |
| week11 | 1.00 | (0.98, 1.02) | 1.02 | (0.98, 1.06) |
| week12 | 1.00 | (0.99, 1.03) | 1.03 | (0.98, 1.07) |
| week13 | 1.01 | (0.99, 1.03) | 1.03 | (0.99, 1.07) |
| week14 | 1.00 | (0.98, 1.02) | 1.03 | (0.99, 1.08) |
| week15 | 1.00 | (0.98, 1.02) | 1.03 | (0.99, 1.08) |
| week16 | 1.01 | (0.98, 1.03) | 1.03 | (0.99, 1.08) |
| week17 | 1.01 | (0.99, 1.03) | 1.03 | (0.99, 1.08) |
| week18 | 1.01 | (0.99, 1.04) | 1.03 | (0.99, 1.08) |
| week19 | 1.01 | (0.99, 1.04) | 1.03 | (0.99, 1.08) |
| week20 | 1.01 | (1.00, 1.04) | 1.03 | (0.99, 1.07) |
| week21 | 1.01 | (0.99, 1.03) | 1.02 | (0.98, 1.06) |
| week22 | 1.01 | (0.99, 1.03) | 1.01 | (0.97, 1.05) |
| week23 | 1.00 | (0.98, 1.02) | 1.00 | (0.96, 1.04) |
| week24 | 1.00 | (0.97, 1.02) | 0.99 | (0.95, 1.03) |
| week25 | 1.00 | (0.98, 1.02) | 1.00 | (0.96, 1.04) |
| week26 | 1.00 | (0.98, 1.02) | 1.00 | (0.96, 1.04) |
| week27 | 1.00 | (0.99, 1.03) | 0.99 | (0.95, 1.03) |
| week28 | 1.00 | (0.99, 1.03) | 0.99 | (0.94, 1.03) |
| week29 | 1.00 | (0.98, 1.02) | 0.99 | (0.94, 1.03) |
| week30 | 1.00 | (0.97, 1.01) | 0.99 | (0.94, 1.04) |
| week31 | 0.99 | (0.96, 1.01) | 0.99 | (0.94, 1.05) |

Effect for ambient temperature was estimated at per 5 °C increase, extreme heat was defined as mean temperature above the $90^{th}$ percentile. Results were derived from the posterior distribution of regression parameters using a logistic-regression distributed lag logistic regression model fitted in the Bayesian setting, adjusted for year of birth, season of conception, birth county, maternal individual characteristics (race/ethnicity, education level, primary insurance type of prenatal care, age at delivery, parity), census-tract level social vulnerability (SES, household, transportation, minority)

**Supplementary Table 3. Characteristics of outcome-discordant siblings in California birth cohort, 2007–2015.**

| | Cerebral Palsy Cases | Controls |
|---|---|---|
| | N=1758 (%) | N=2316 (%) |
| Maternal age at delivery | | |
| <=18 | 71 (4.0%) | 65 (2.8%) |
| 19-25 | 478 (27.2%) | 674 (29.1%) |
| 26-30 | 520 (29.6%) | 661 (28.5%) |
| 31-35 | 451 (25.7%) | 560 (24.2%) |
| >35 | 238 (13.5%) | 356 (15.4%) |
| Maternal race and ethnicity | | |
| Non-Hispanic White | 514 (29.2%) | 680 (29.4%) |
| Hispanic or Latinx of any race | 942 (53.6%) | 1,238 (53.5%) |
| African American or Black | 84 (4.8%) | 116 (5.0%) |
| Asian | 164 (9.3%) | 212 (9.2%) |
| Others (include Pacific Islanders and other) | 18 (1.0%) | 32 (1.4%) |
| Unknown | 36 (2.0%) | 38 (1.6%) |
| Maternal education level | | |
| <12th grade | 439 (25.0%) | 581 (25.1%) |
| High school or diploma | 835 (47.5%) | 1,109 (47.9%) |
| College and above | 411 (23.4%) | 535 (23.1%) |
| Unknown | 73 (4.2%) | 91 (3.9%) |
| Primary payment type for prenatal care | | |
| Government | 833 (47.4%) | 1191 (51.4%) |
| Private | 860 (48.9%) | 1067 (46.1%) |
| Other | 57 (3.3%) | 54 (2.4%) |
| Unknown | 8 (0.5%) | 4 (0.2%) |
| Sex of children | | |
| Male | 965 (54.9%) | 1,193 (51.5%) |
| Female | 793 (45.1%) | 1,123 (48.5%) |
| Parity | | |
| 1 | 564 (32.1%) | 593 (25.6%) |
| 2 | 613 (34.9%) | 807 (34.8%) |
| 3+ | 581 (33.0%) | 916 (39.6%) |
| Birth year | | |
| 2005-2010 | 3631 (61.2%) | 613,831 (56.2%) |
| 2011-2015 | 2307 (38.9%) | 478,482 (43.8%) |
| Season of conception | | |
| Spring (March-May) | 432 (24.6%) | 571 (24.7%) |
| Summer (June-August) | 440 (25.0%) | 525 (22.7%) |
| Fall (September-November) | 411 (23.4%) | 588 (25.4%) |
| Winter (December-February) | 416 (23.7%) | 562 (24.3%) |
| Unknown | 59 (3.4%) | 70 (3.0%) |
| Gestational length (week) (mean ± SD) | 37.5 ± 6.5 | 38.8 ± 4.7 |
| Social vulnerability index (mean ± SD) | | |
| Total score | 0.61 ± 0.28 | 0.61 ± 0.28 |
| Socioeconomic domain | 0.61 ± 0.27 | 0.61 ± 0.27 |
| Household domain | 0.59 ± 0.28 | 0.59 ± 0.28 |
| Minority domain | 0.59 ± 0.28 | 0.59 ± 0.28 |
| Transportation domain | 0.57 ± 0.28 | 0.57 ± 0.28 |

**Supplementary Table 4. Estimated odds ratio (ORs) and 95% credible intervals (CIs) for the cerebral palsy exposed to ambient temperature and extreme heat in gestational week 0 to 31 among siblings (match Figure 2).**

| Gestational week | Ambient Temperature OR (95% CI) | Extreme Heat OR (95% CI) |
|---|---|---|
| Week 0 | 1.02 (0.95, 1.09) | 1.03 (0.79, 1.34) |
| Week 1 | 1.05 (0.98, 1.12) | 1.10 (0.84, 1.44) |
| Week 2 | 1.08 (1.01, 1.16) | 1.25 (0.97, 1.61) |
| Week 3 | 1.06 (0.99, 1.13) | 1.36 (1.04, 1.79) |
| Week 4 | 1.06 (0.99, 1.13) | 1.63 (1.24, 2.13) |
| Week 5 | 1.07 (1.00, 1.15) | 1.28 (0.97, 1.69) |
| Week 6 | 1.09 (1.01, 1.16) | 1.42 (1.07, 1.90) |
| Week 7 | 1.07 (1.00, 1.15) | 1.23 (0.96, 1.59) |
| Week 8 | 1.04 (0.97, 1.11) | 1.15 (0.88, 1.50) |
| Week 9 | 1.07 (1.00, 1.14) | 1.27 (0.98, 1.65) |
| Week 10 | 1.04 (0.97, 1.11) | 1.03 (0.79, 1.35) |
| Week 11 | 1.03 (0.97, 1.11) | 0.89 (0.68, 1.16) |
| Week 12 | 1.05 (0.98, 1.13) | 1.15 (0.88, 1.49) |
| Week 13 | 1.04 (0.97, 1.12) | 1.10 (0.83, 1.45) |
| Week 14 | 1.01 (0.94, 1.08) | 0.90 (0.69, 1.17) |
| Week 15 | 0.98 (0.91, 1.05) | 0.88 (0.67, 1.14) |
| Week 16 | 0.99 (0.93, 1.06) | 1.03 (0.79, 1.33) |
| Week 17 | 0.98 (0.92, 1.05) | 0.74 (0.56, 0.97) |
| Week 18 | 0.98 (0.92, 1.05) | 0.86 (0.67, 1.11) |
| Week 19 | 0.96 (0.89, 1.02) | 0.81 (0.63, 1.05) |
| Week 20 | 0.96 (0.90, 1.03) | 0.95 (0.73, 1.23) |
| Week 21 | 0.92 (0.86, 0.99) | 0.80 (0.62, 1.04) |
| Week 22 | 0.95 (0.89, 1.02) | 0.93 (0.71, 1.20) |
| Week 23 | 0.95 (0.89, 1.01) | 0.90 (0.70, 1.16) |
| Week 24 | 0.93 (0.86, 0.99) | 0.90 (0.69, 1.16) |
| Week 25 | 0.93 (0.87, 1.00) | 1.02 (0.78, 1.33) |
| Week 26 | 0.93 (0.87, 1.00) | 1.03 (0.79, 1.34) |
| Week 27 | 0.92 (0.86, 0.99) | 0.92 (0.71, 1.20) |
| Week 28 | 0.93 (0.87, 1.00) | 0.93 (0.71, 1.22) |
| Week 29 | 0.93 (0.86, 1.00) | 1.06 (0.81, 1.38) |
| Week 30 | 0.91 (0.84, 0.98) | 0.81 (0.61, 1.07) |
| Week 31 | 0.92 (0.86, 0.99) | 0.91 (0.68, 1.20) |

Effect for ambient temperature was estimated at per 5 °C increase, extreme heat was defined as mean temperature above the 90th percentile. Results were derived from the conditional logistic regression applied separately to each week, adjusted for year of birth, maternal primary insurance type of prenatal care, age at delivery, and parity.

**Supplementary Figure 1. Estimated odds ratio (ORs) and 95% credible intervals (CIs) for the cerebral palsy exposed to ambient temperature and extreme heat in gestational week 0 to 36 during pregnancy.**

**Ambient temperature**

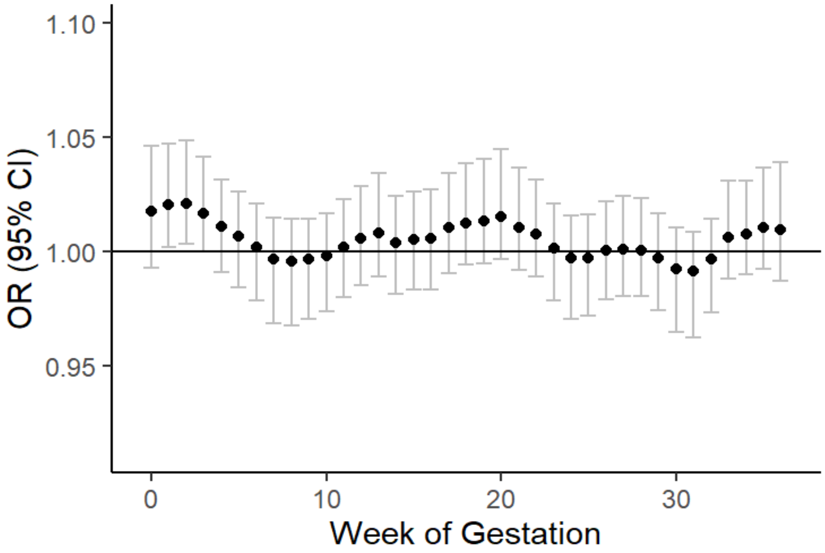


**Extreme hot**

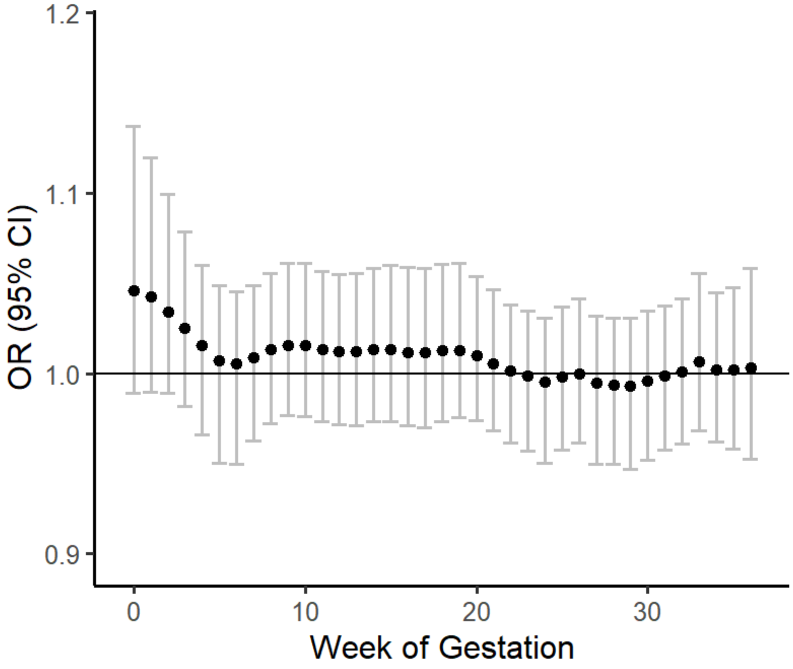

**Supplementary Figure 2. Estimated odds ratio (ORs) and 95% credible intervals (CIs) for the cerebral palsy exposed to ambient temperature and extreme heat in the final seven weeks preceding birth.**

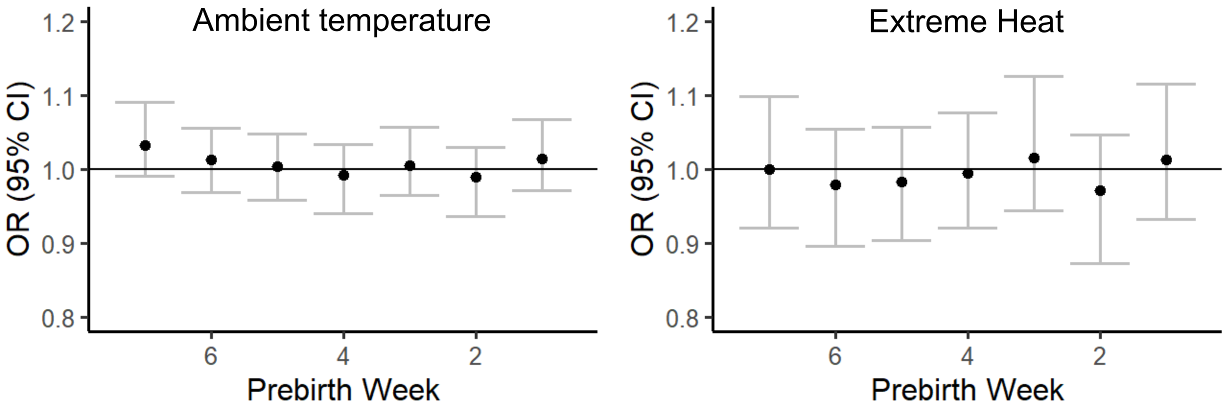

**Supplementary Figure 3. Cumulative Odds Ratios (ORs) and 95% CIs for cerebral palsy according to exposures to ambient temperature and extreme heat in final seven weeks preceding birth, stratified by length of gestation**

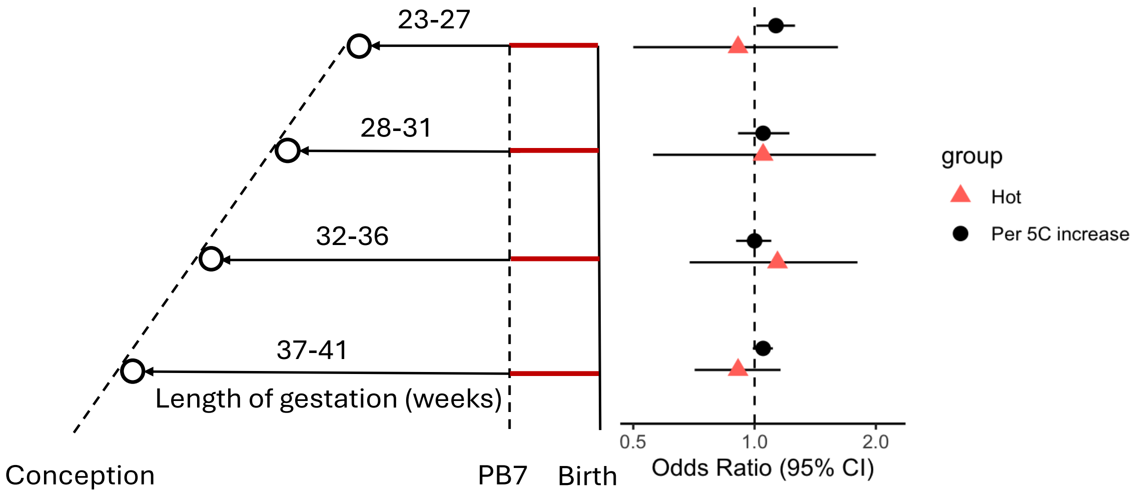

**Supplementary Figure 3. Susceptible window for the cerebral palsy exposed to ambient temperature and extreme heat in gestational week 0 to 31 during pregnancy, stratified by cerebral palsy subtypes.**

**Ambient Temperature**

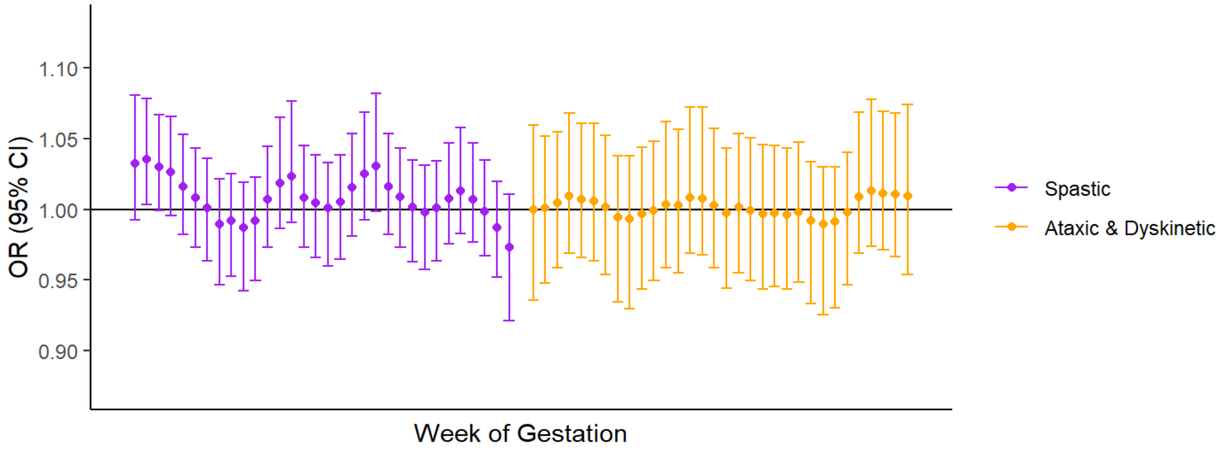


**Extreme heat**

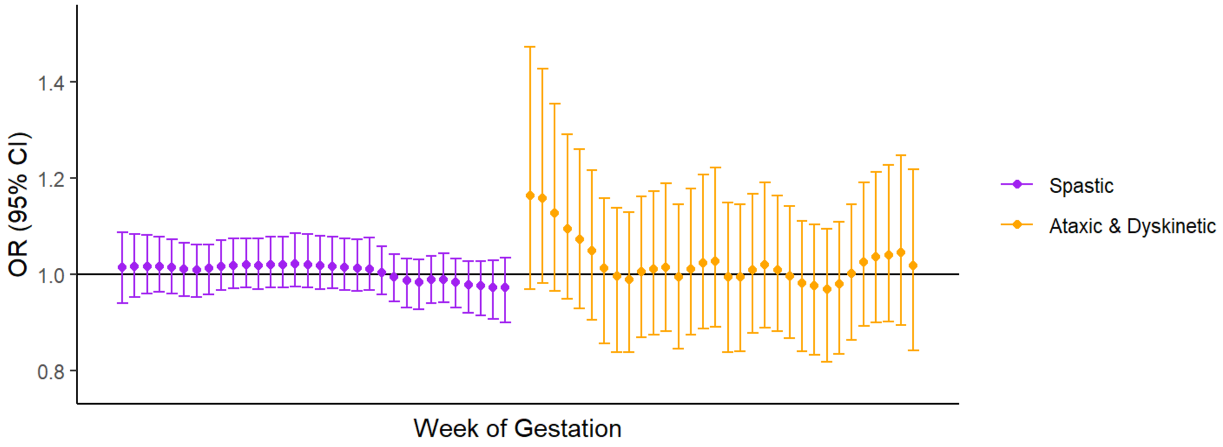

**Supplementary Figure 4. Susceptible window for the cerebral palsy exposed to ambient temperature and extreme heat in gestational week 0 to 31 during pregnancy, stratified by child's sex.**

**Ambient Temperature**

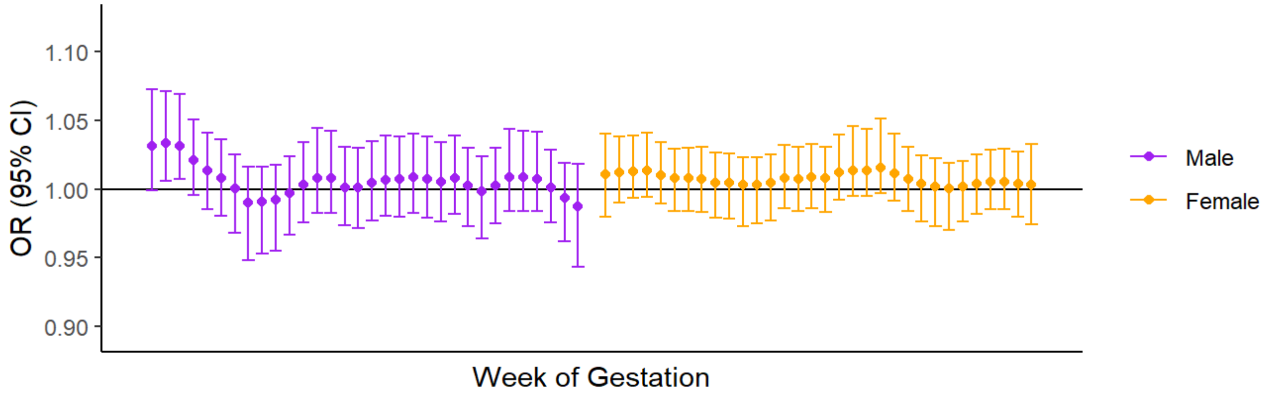


**Extreme heat**

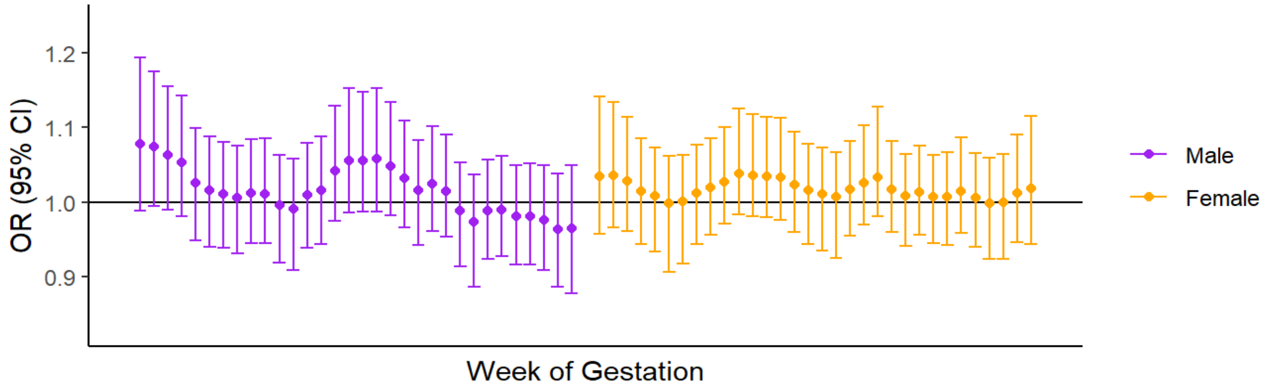

**Supplementary Figure 5. Susceptible window for the cerebral palsy exposed to ambient temperature and extreme heat in gestational week 0 to 31 during pregnancy, stratified by maternal race/ethnicity.**

### Ambient Temperature

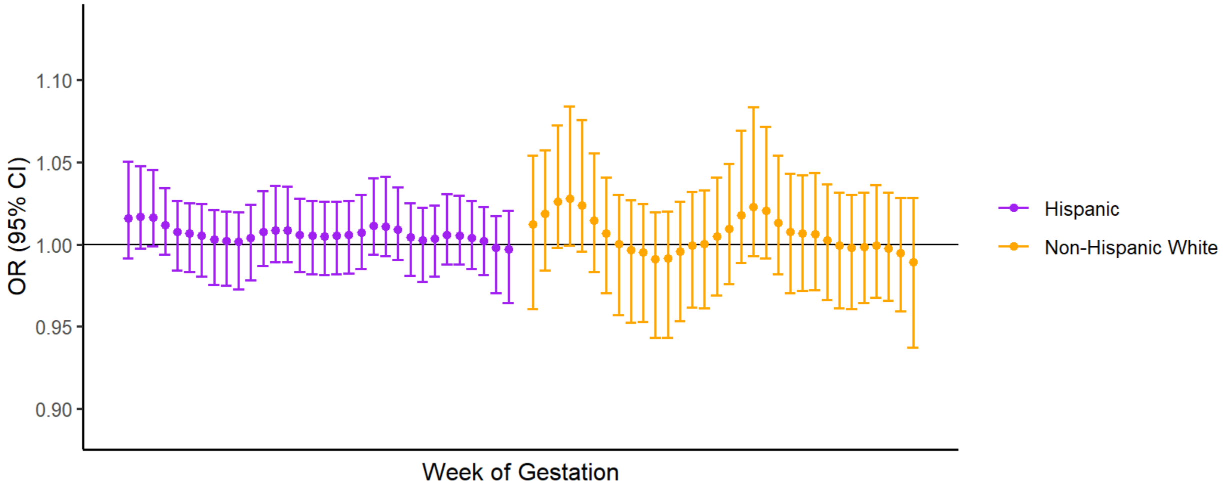


### Extreme heat

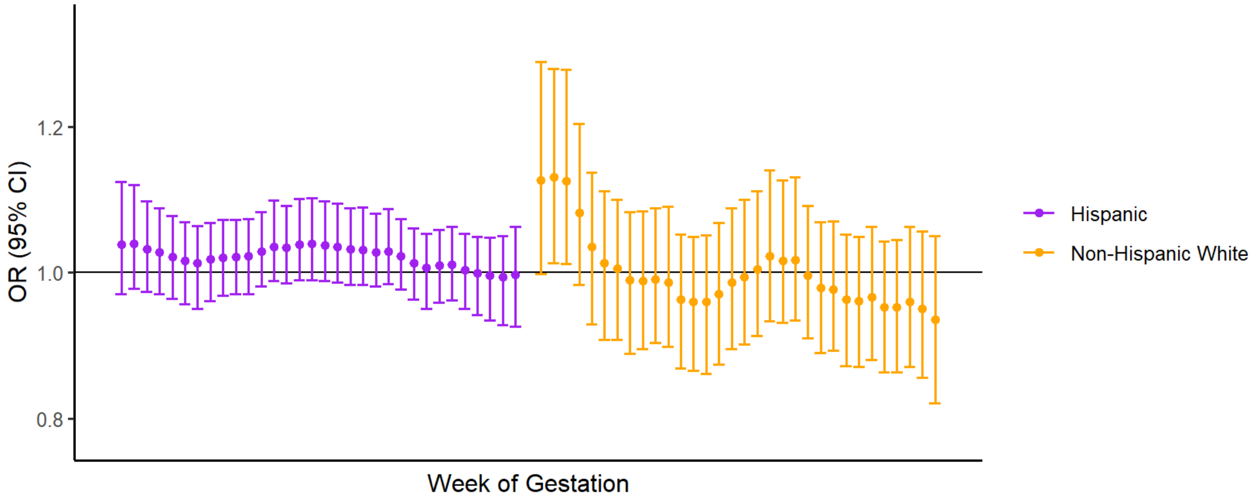

**Supplementary Figure 6. Susceptible window for the cerebral palsy exposed to ambient temperature and extreme heat in gestational week 0 to 31 during pregnancy, stratified by maternal education level.**

### Ambient Temperature

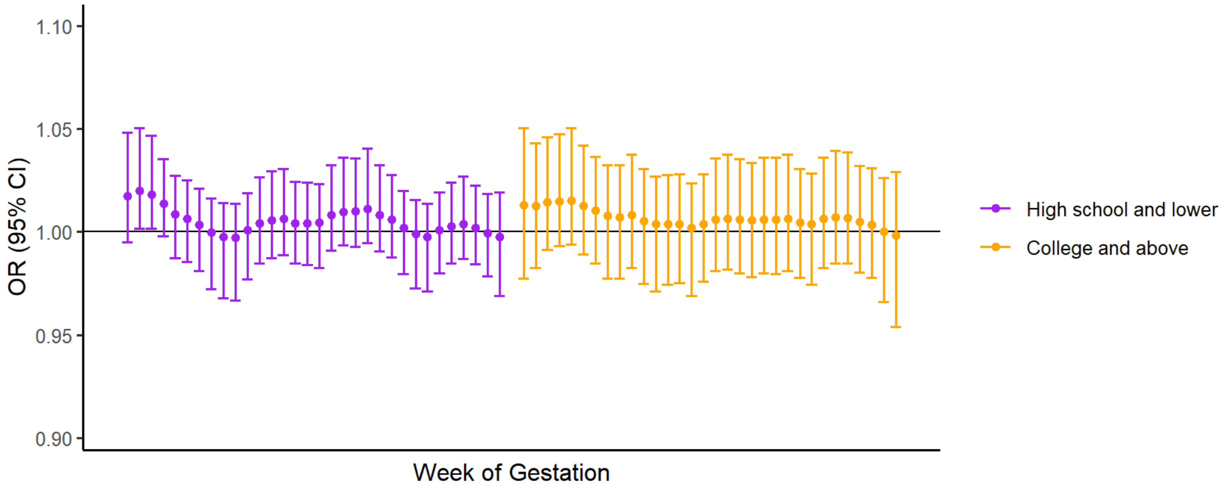


### Extreme heat

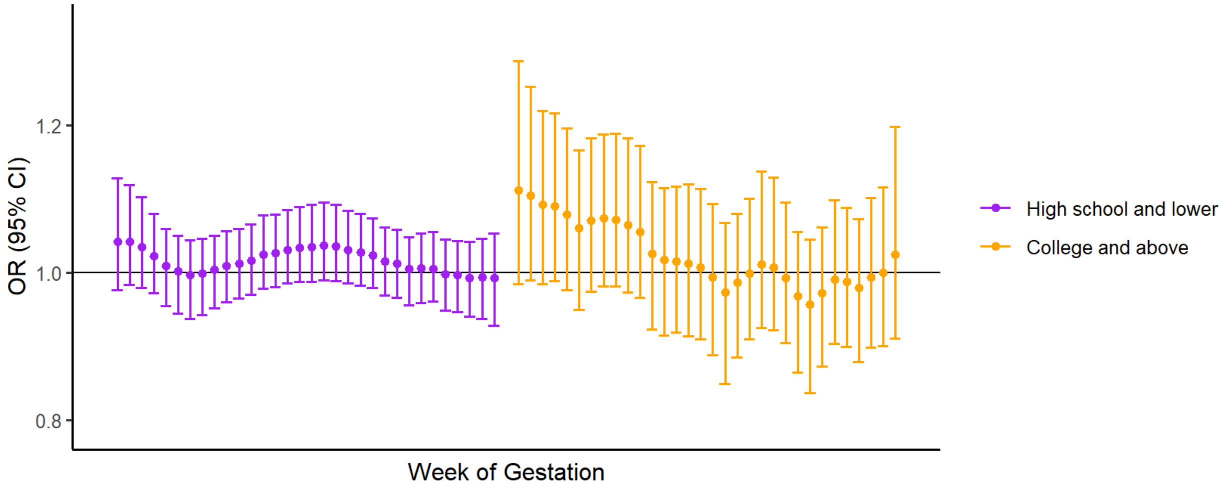

**Supplementary Figure 8. Susceptible window for the cerebral palsy exposed to ambient temperature and extreme heat in gestational week 0 to 31 during pregnancy, stratified by census-tract level social vulnerability index.**

**Ambient Temperature**

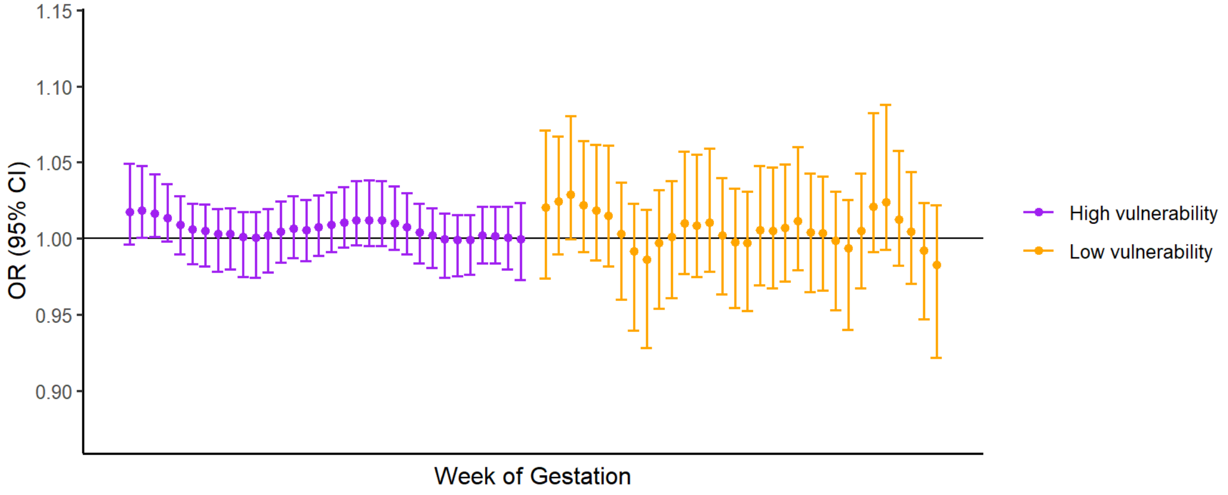


**Extreme heat**

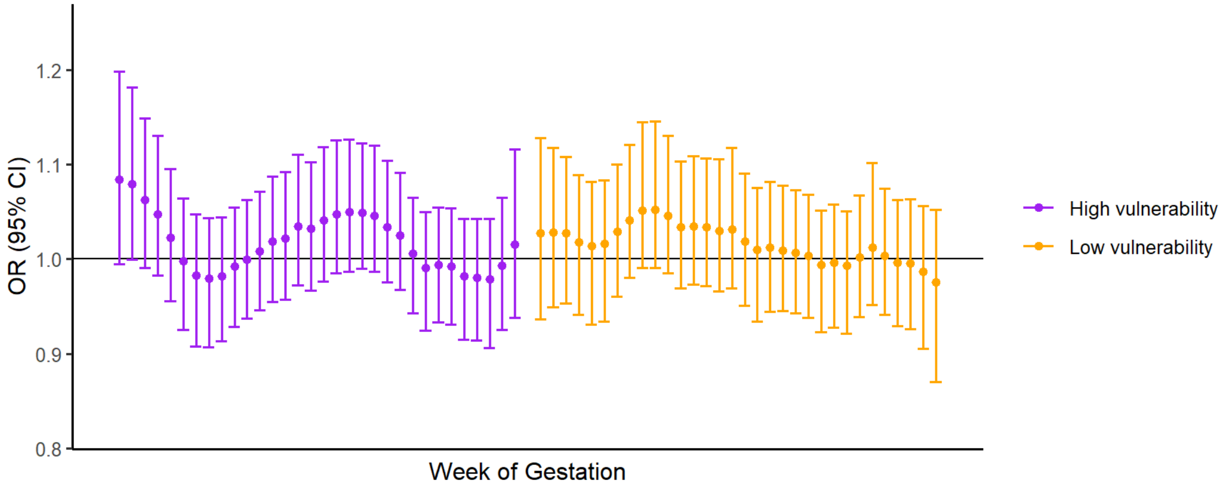

**Supplementary Figure 9. Non-linear relationship of ambient temperature and cerebral palsy in gestational week 0 to 3, with degree freedoms of 3 to 6**

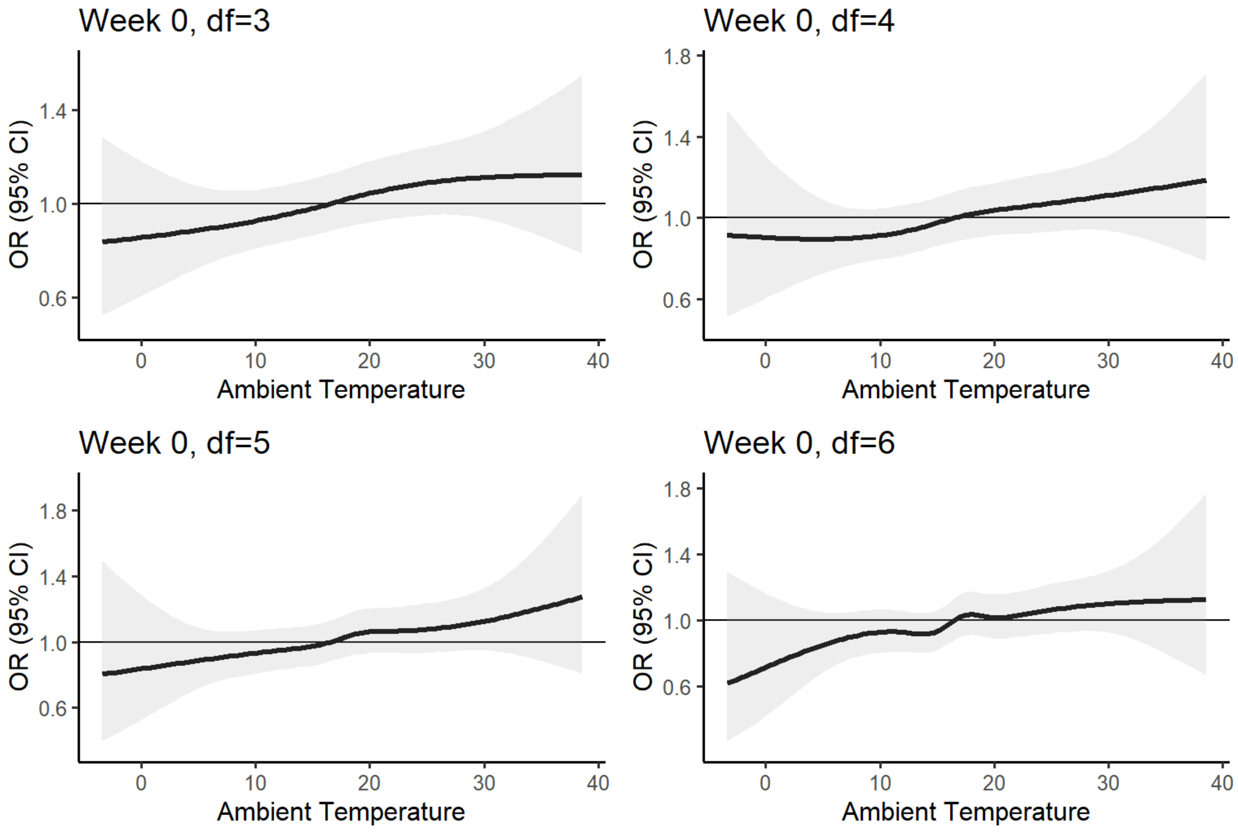

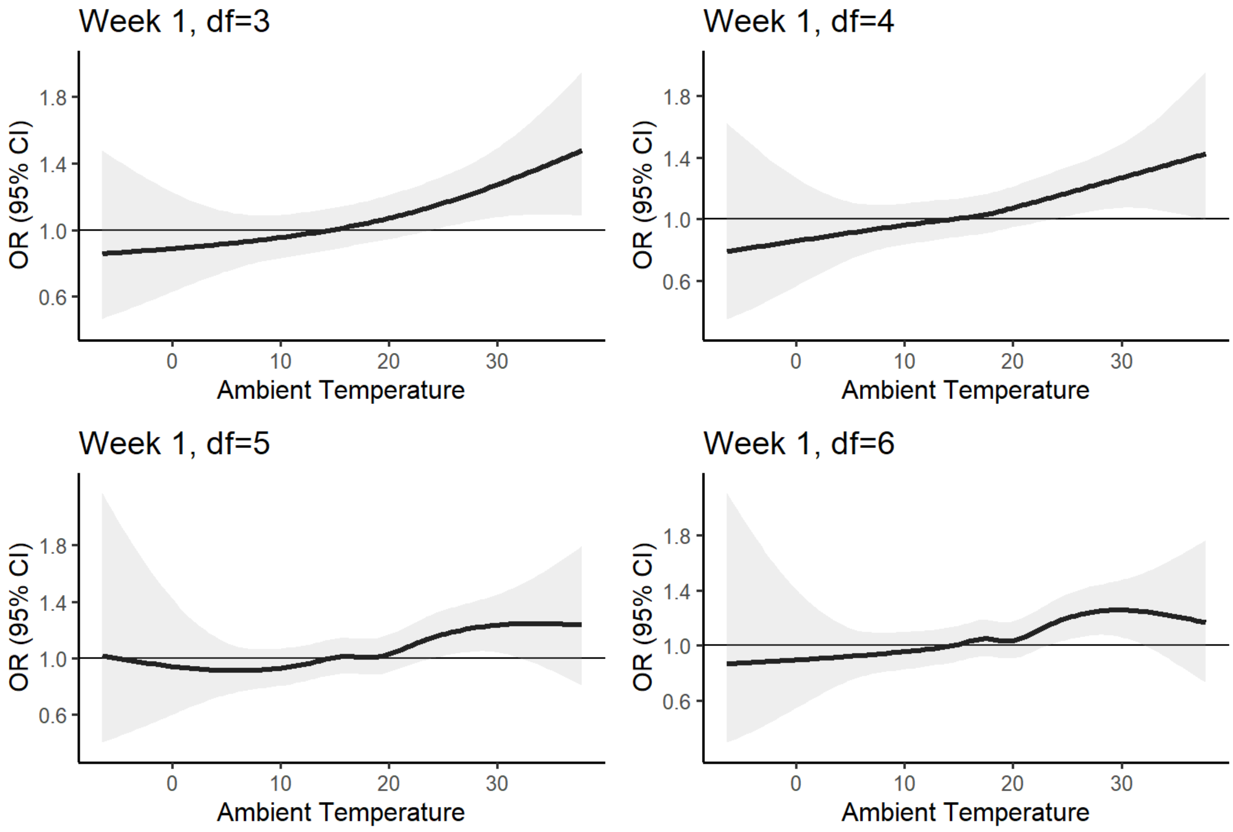

Week 1, df=3
OR (95% CI)
1.8
1.4
1.0
0.6
0
10
20
30
Ambient Temperature
Week 1, df=4
OR (95% CI)
1.8
1.4
1.0
0.6
0
10
20
30
Ambient Temperature
Week 1, df=5
OR (95% CI)
1.8
1.4
1.0
0.6
0
10
20
30
Ambient Temperature
Week 1, df=6
OR (95% CI)
1.8
1.4
1.0
0.6
0
10
20
30
Ambient Temperature

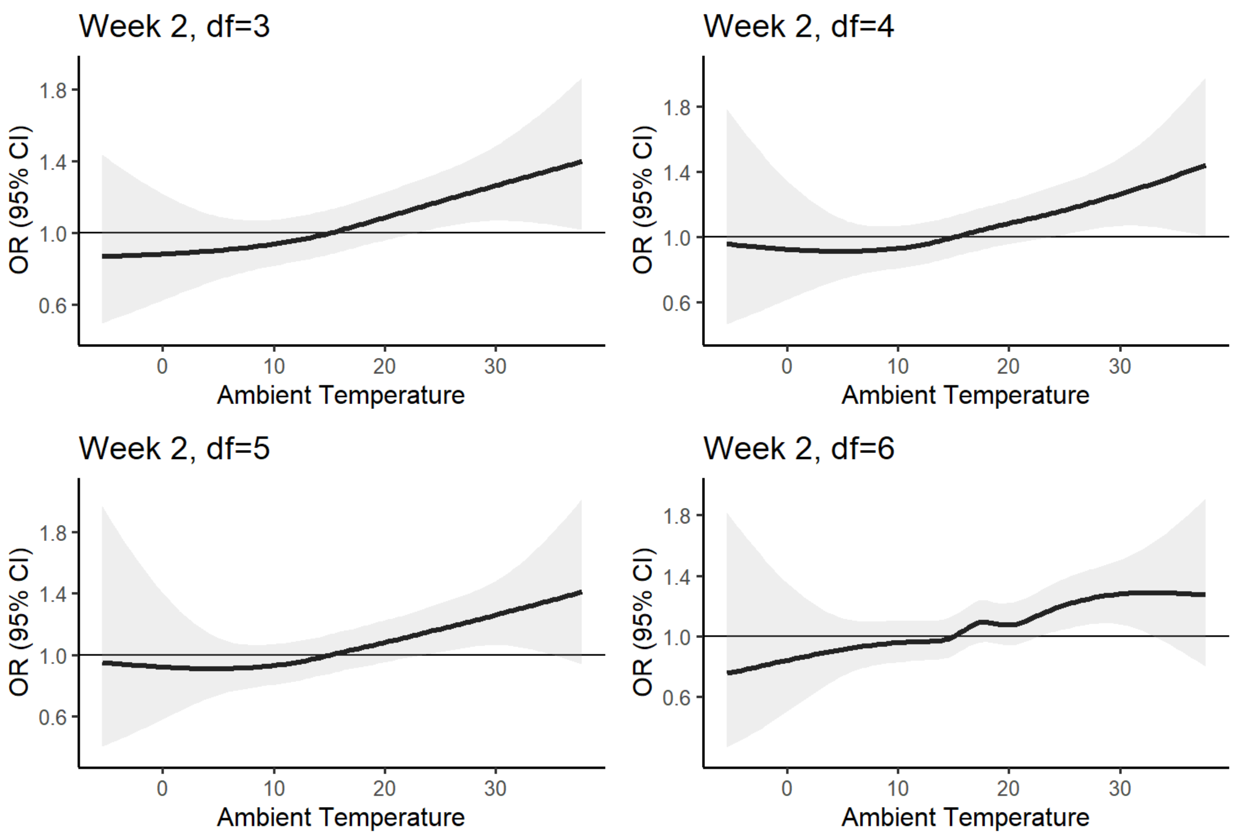

Week 2, df=3
Week 2, df=4
Week 2, df=5
Week 2, df=6
OR (95% CI)
1.8
1.4
1.0
0.6
0
10
20
30
Ambient Temperature

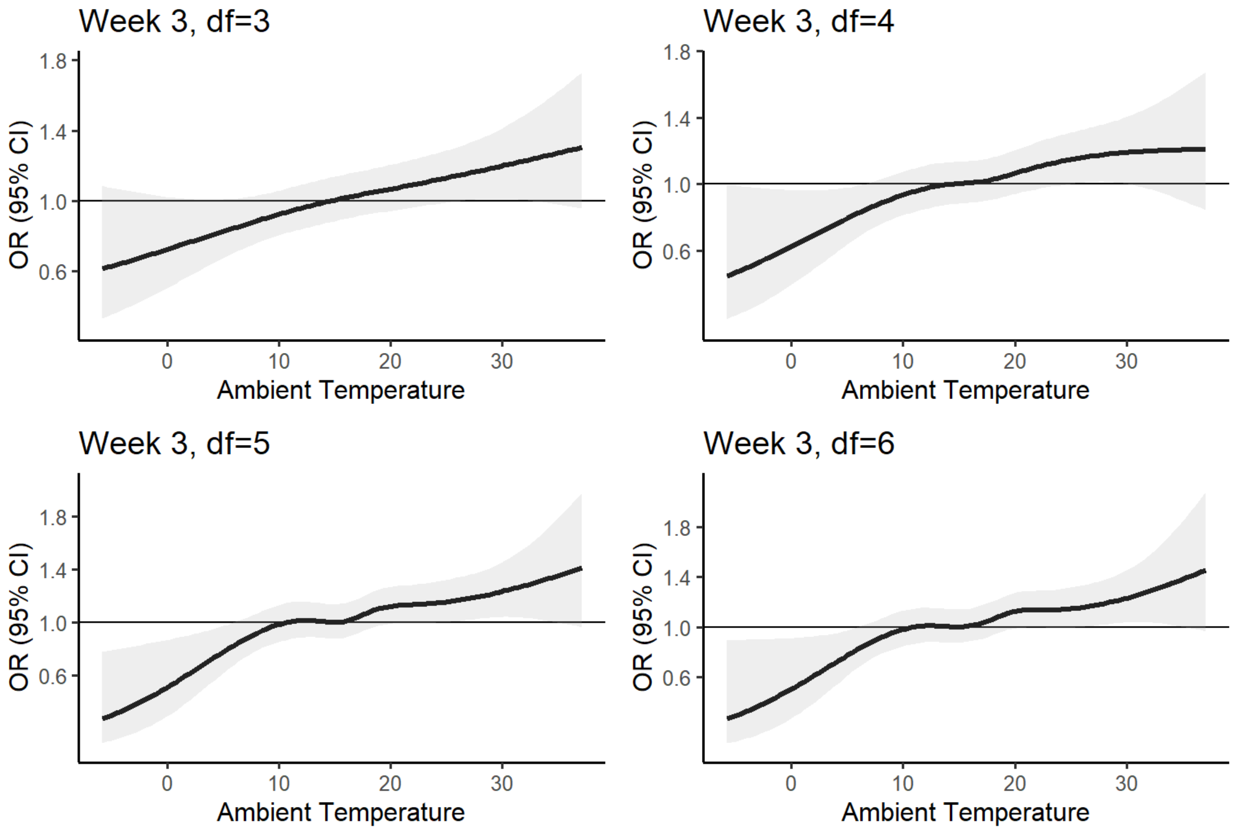

Week 3, df=3
Week 3, df=4
Week 3, df=5
Week 3, df=6
OR (95% CI)
Ambient Temperature
1.8
1.4
1.0
0.6
0
10
20
30

**Supplementary Figure 10. Underlying biological pathways from ambient temperature exposure and cerebral palsy**

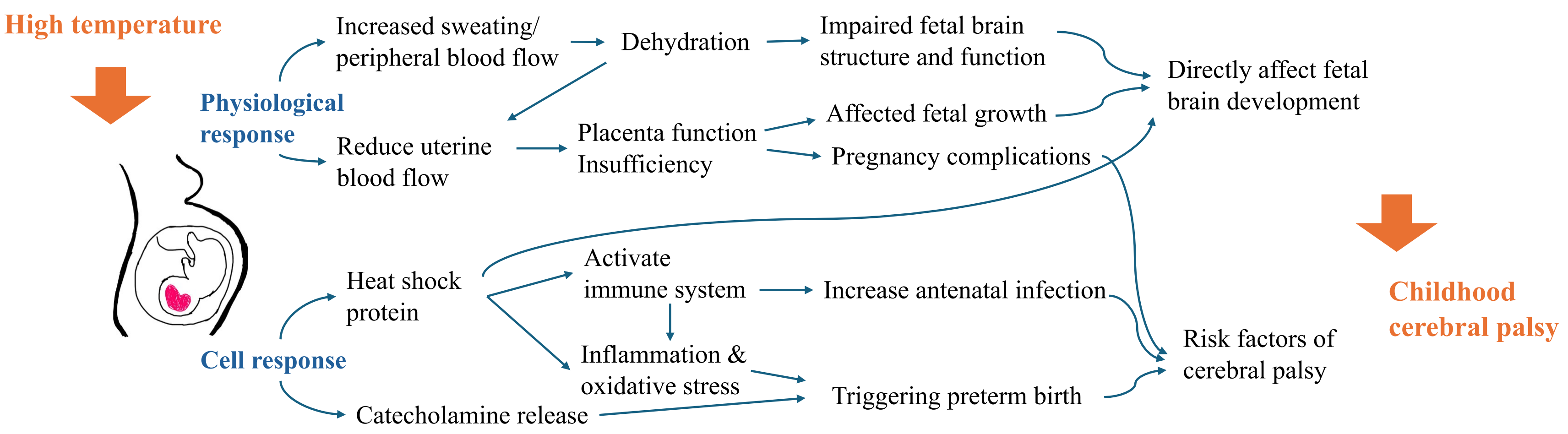


Plot Reference:
Samuels, et al. *Int J Biomet*, 2022; Ha. *Curr Environ Health Rep*, 2022;; Dubrez L, et al. *Oncogene*. 2020; Kiyatkin EA. *Temperature*. 2019; Ponomarenko M, et al. *Academic Press*2013.
